# International Randomised Controlled Trial evaluating metabolic syndrome in type 2 Diabetic Cigarette Smokers following switching to Combustion-Free Nicotine Delivery Systems: the DIASMOKE protocol

**DOI:** 10.1101/2020.08.19.20177253

**Authors:** Arkadiusz Krysiński, Cristina Russo, Sarah John, Jonathan Belsey, Davide Campagna, Pasquale Caponnetto, Lorina Vudu, Chong Wei Lim, Francesco Purrello, Maurizio Di Mauro, Farrukh Iqbal, David Fluck, Edward Franek, Riccardo Polosa, Pankaj Sharma, on behalf of the DIASMOKE collaborators

## Abstract

Reducing exposure to cigarette smoke is an imperative for public health, and even more so for diabetic patients. Increasingly, combustion-free technologies for nicotine delivery such as e-cigarettes and heated tobacco products are substituting conventional cigarettes and accelerating the current downward trends in smoking prevalence. However, there is limited information about the long-term health impact in diabetics who use these technologies.

This international randomised controlled trial of type 2 diabetic cigarette smokers will test the hypothesis that following a switch from conventional cigarettes to Combustion-Free Nicotine Delivery Systems (C-F NDS), a measurable improvement in metabolic syndrome (MetS) risk factors and functional parameters will be shown over the course of 2 years.

A total of 576 patients will be randomized (1:2 ratio) to either a control arm (Study Arm A), in which they will be offered referral to smoking cessation programs or to an intervention arm (Study Arm B), in which they will be assigned to C-F NDS use. The primary outcome will be the prevalence of the a difference in MetS score between baseline and 2 years follow-up, with comparison being made between diabetic patients randomized to each arm of the study. Patient recruitment will start in October 2020 and enrolment is expected to be completed by August 2021.

This will be the first study determining the overall health impact of using such technologies in diabetic patients. Data from this study will provide valuable insights into the overall potential of C-F NDS to reduce the risk of cardiovascular disease in individuals, particularly diabetic patients.

Clinical Trial Registration: https://clinicaltrials.gov/ct2/show/NCT04231838

## BACKGROUND

Diabetes mellitus (DM) can cause irreversible damage to the blood vessels leading to microvascular (retinopathy, nephropathy and diabetic neuropathy) or macrovascular (coronary artery disease, stroke, peripheral arterial disease) complications (1), the latter cardiovascular complications being most common, and a frequent cause of death. Besides diabetes and hyperglycemia, obesity, hypertension, dyslipidemia are well established cardiovascular risk factors, all of which come under the umbrella definition of metabolic syndrome (MetS). Other cardiovascular risk factors may also coexist in these patients, the most important being smoking.

Cigarette smoking is a strong cardiovascular risk factor not included in the definition of MetS but substantially increases the risk of micro- and macrovascular complications in patients with type 2 DM (T2DM) (2–5), whereas quitting smoking substantially reduces this risk (4–7). Given that exposure to cigarette smoke is associated with vascular damage, endothelial dysfunction and activation of coagulation and fibrinolysis (8), it is not surprising that smoking enhances the combined harmful effects of elevated blood glucose and other risk factors and accelerates vascular damage in diabetic patients. If reducing exposure to cigarette smoke is an imperative for public health, it is even more so for patients with T2DM (9). However, prevalence of smoking among people with DM appears to be similar to that of the general population (10). In the United States, the prevalence of tobacco consumption has decreased substantially, but this beneficial trend has not been observed in patients with DM (11).

There is a clear urgent need to target T2DM patients to successful smoking cessation therapies, such as nicotine-containing preparations (12,13). Unfortunately, there is no convincing demonstration of effective cessation interventions in patients with diabetes (14) and, in general, most smokers are reluctant to seek formal treatment for stopping smoking with the vast majority making attempts to quit without assistance (15,16). Consequently, the need for novel and more efficient approaches is required.

Combustion-free technologies for nicotine delivery such as e-cigarettes (ECs) and heated tobacco products (HTPs) are substituting conventional cigarettes globally (17) and are thought to be less harmful alternative to tobacco smoking (18–20). However, there are no long-term studies assessing cardiovasular risk or effect on cardiovascular risk factors in diabetics who use these technologies.

The DIASMOKE collaborators seek to determine whether T2DM cigarette smokers who switch to combustion-free nicotine delivery systems (C-F NDS) experience measurable improvements in their cardiovascular risk parameters.

## METHODS

DIASMOKE (Assessing the impact of combustion free-nicotine delivery technologies in DIAbetic SMOKErs) is an international, multicentre, open label randomized controlled study designed to determine whether T2DM cigarette smokers switching to C-F NDS experience measurable improvement in cardiovascular risk parameters as a consequence of avoiding exposure to cigarette smoke toxicants.

### Study Population

The inclusion and exclusion criteria are summarized in **Table 1**. Participants will be cigarette smokers with a clinical diagnosis of T2DM. They will be a minimum of 23 years of age, any gender, and with a body mass index (BMI) between 17.6 and 32.0 kg/m2 (inclusive), body weight exceeding 50 kg (males) or 40 kg (females), and HbA1C values between 6.5% and 10%.

**Table 1:**
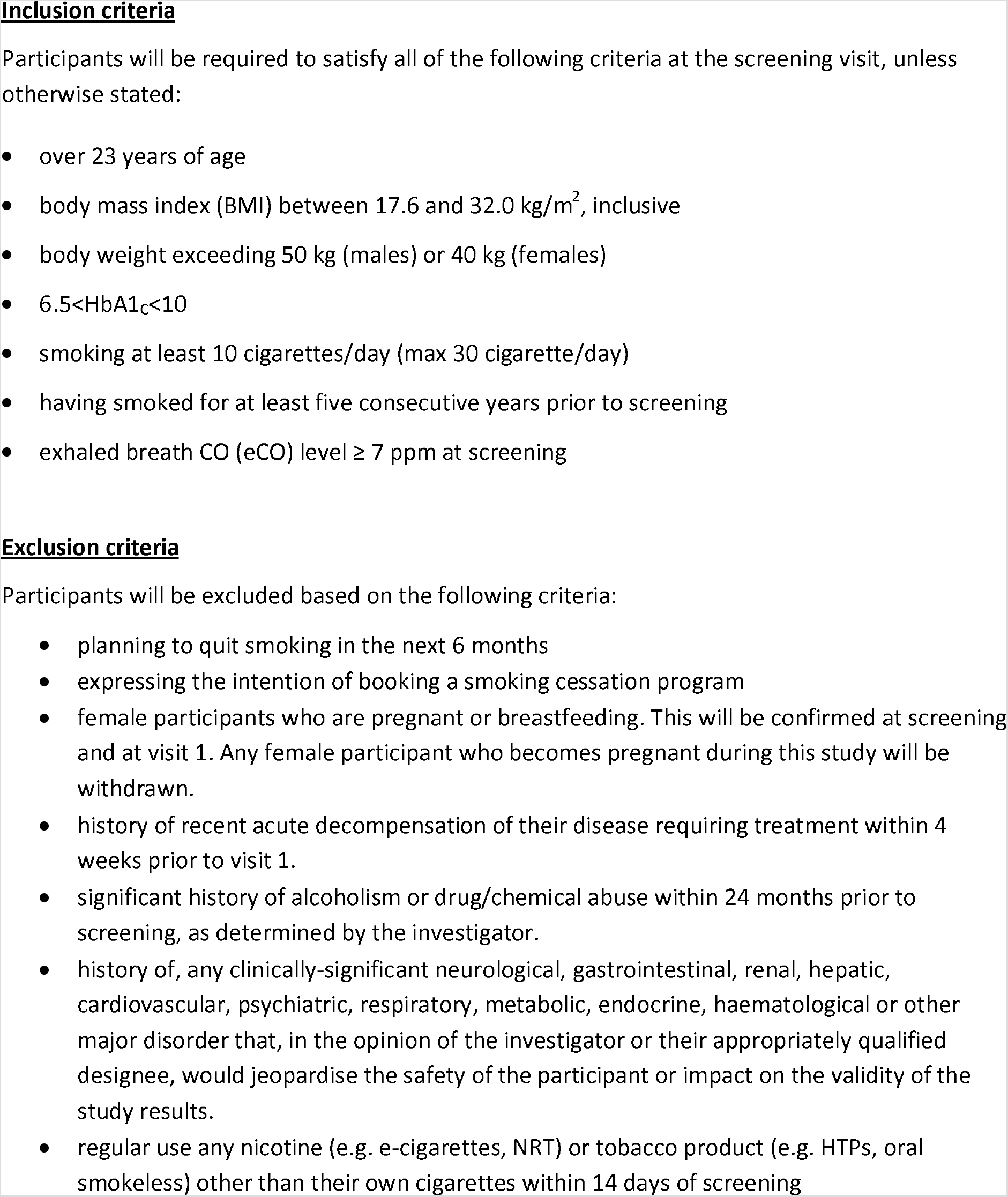
PARTICIPANTS ELIGIBILITY CRITERIA.

Only regular cigarette smokers (>10/day for at least 5 years) will be considered for inclusion. Smoking status will be verified by an exhaled CO measurement (exhaled CO ≥ 7 ppm). Each participant will be offered access to local free smoking cessation programs, and only those that refuse participation in cessation programs and are willing to switch to a C-F NDS will be randomized following informed consent.

### Ethical Approval

The study will be conducted according to the Principles of Good Clinical Practice (GCP) and Declaration of Helsinki. All six local Ethics Committees reviewed and approved the study and - where appropriate - translated relevant documentation (informed consent form, patients information sheet, etc). The trial is registered with ClinicalTrails.gov Identifier: NCT04231838.

### Study Design

The study design flow of DIASMOKE is illustrated in **Figure 1**. Participants will attend a Screening Visit within 28 days prior to Visit 1 and undergo demographic assessments including socio-demographic data, detailed medical history (including medication use), detailed smoking, vaping, and heated tobacco products use history and their intention to quit. Modification in their diet and/or anti-diabetic medication will be recorded regularly throughout the study. All patients will be offered smoking cessation program as per local guidelines. Participants will be offered a further second opportunity to enroll in the free local smoking cessation program prior to enrollment.

**Figure 1:**
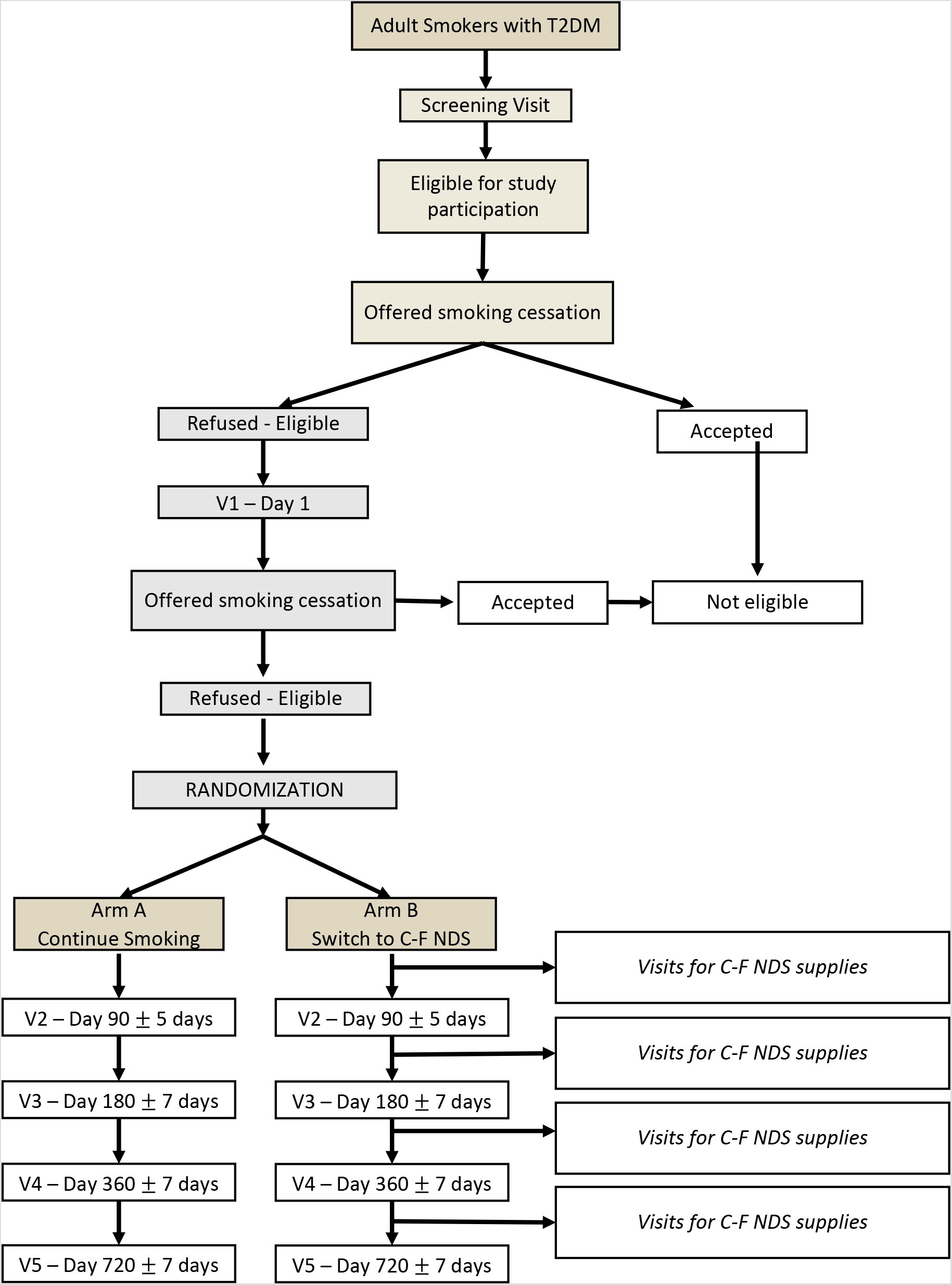
STUDY DESIGN FLOW OF DIASMOKE

Following baseline assessments on Day 1 (see **Table 2a,2b)**, participants will be randomized to either control (A) or the intervention (B) arms in ratio of 1:2 (arm A: arm B) in order to compensate for the estimated 50% drop-out rate. Patients randomized into arm B will be allowed to choose the product of their preference from the given pool of most popular C-F NDS. The participants will be trained and counselled on the chosen device and given a full one week supply of tobacco sticks/e-cigarette cartridges/e-liquids refill bottles prior to check-out on Day 1. After randomization, a dedicated tracker APP will be installed on patients’ smartphones. The APP is designed to track patients behavior (physical activity, adherence to sugar testing, cigarette smoking frequency, daily C-F NDS usage) to identify protocol violations that will generate flagging events and alerts, to collect adverse events and to send reminders (next scheduled appointment, study restrictions, instructions, etc) throughout the whole duration of the study.

Subsequently, participants will be invited to attend four further clinical visits conducted in an ambulatory setting (Visits 2-5) to undergo a range of measurements and blood tests (see **Table 2a,2b)**. Following each visit participants will be supplied with an appropriate amount of consumables (tobacco sticks, e-cigarette cartridges, e-liquid refill bottles). Participants will fast overnight (from midnight) prior to each study visit at which clinical laboratory evaluations will be performed. Patients will be instructed to refrain from consuming alcohol for 24 hours prior to clinic visits and instructed not to consume more than 14 units of alcohol per week for the entire duration of the study.

**Table 2a:** STUDY ASSESSMENTS (Arm A – continue smoking)

| **Procedure** | **Screen**<br>**(V0)**<br>**<28days before V1** | **Baseline**<br>**(V1)**<br>**Day 1** | **Week12**<br>**(V2)**<br>**Day 90 ±5** | **Week24**<br>**(V3)**<br>**Day 180 ±7** | **Week52**<br>**(V4)**<br>**Day 360 ± 7** | **Week104**<br>**(V5)**<br>**Day 720 ±7** |
| --- | --- | --- | --- | --- | --- | --- |
| Eligibility Criteria check | X | X |  |  |  |  |
| Medical Hx | X |  |  |  |  |  |
| Antidiabetic drug use |  | X | X | X | X | X |
| Smoking / Vaping Hx | X |  |  |  |  |  |
| Cigarette consumption check | X | X | X | X | X | X |
| Exhaled CO |  | X | X | X | X | X |
| FTND |  | X |  |  |  |  |
| Intention to quit | X | X |  |  |  |  |
| Informed Consent Forms |  | X |  |  |  |  |
| Randomization |  | X |  |  |  |  |
| Tracker APP installation |  | X |  |  |  |  |
| Vital Signs |  | X | X | X | X | X |
| Weight, height, BMI, waist circumference |  | X | X | X | X | X |
| Body composition equation* |  | X | X | X | X | X |
| Diabetes QoL questionnaire |  | X | X | X | X | X |
| Smoking Cessation Advice | X | X | X | X | X | X |
| Spirometry |  | X |  |  |  | X |
| Metabolic Syndrome (NCEP) assessment** |  | X | X | X | X | X |
| Fasting blood sample to measure: CBC (WBC Hb, platelets), lipid profile (Triglycerides, LDL and HDL cholesterol) HbA1c, Insulin level |  | X | X | X | X | X |
| Testosterone (men only) |  | X | X | X | X | X |

| **Procedure** | **Screen**<br>**(V0)**<br><28days before V1 | **Baseline**<br>**(V1)**<br>**Day 1** | **Week12**<br>**(V2)**<br>Day 90 ±5 | **Week24**<br>**(V3)**<br>Day 180 ±7 | **Week52**<br>**(V4)**<br>Day 360 ± 7 | **Week104**<br>**(V5)**<br>Day 720 ±7 |
| --- | --- | --- | --- | --- | --- | --- |
| Urine Albumin to Creatinine Ratio (ACR) |  |  |  |  |  |  |
| Safety reporting (AE and SAE)/e-CRF |  | **X** | **X** | **X** | **X** | **X** |

\*Body composition (fat and skeletal muscle mass) by validated equation by Lean ME et al. (1996) and Al-Gindan YY et al. (2014)
\*\*The Metabolic Syndrome is defined according to the National Cholesterol Education Program (NCEP) criteria as having $\geq 3$ of the following five features:
1-Fasting plasma glucose $\geq 6.1$ mmol/L (110 mg/dL)
2- Blood pressure $\geq 130/\geq 85$ mmHg or treatment for hypertension
3-Triglycerides $\geq 1.69$ mmol/L (150 mg/dL) or treatment for high triglycerides
4-High-density lipoprotein (HDL) cholesterol $<1.29$ mmol/L (50 mg/dL) for females or $<1.04$ mmol/L (40 mg/dL) for males or treatment for low HDL
5-Waist circumference $\geq 88$ cm (35 in) for females or $\geq 102$ cm (40 in) for males.

**Table 2b:**
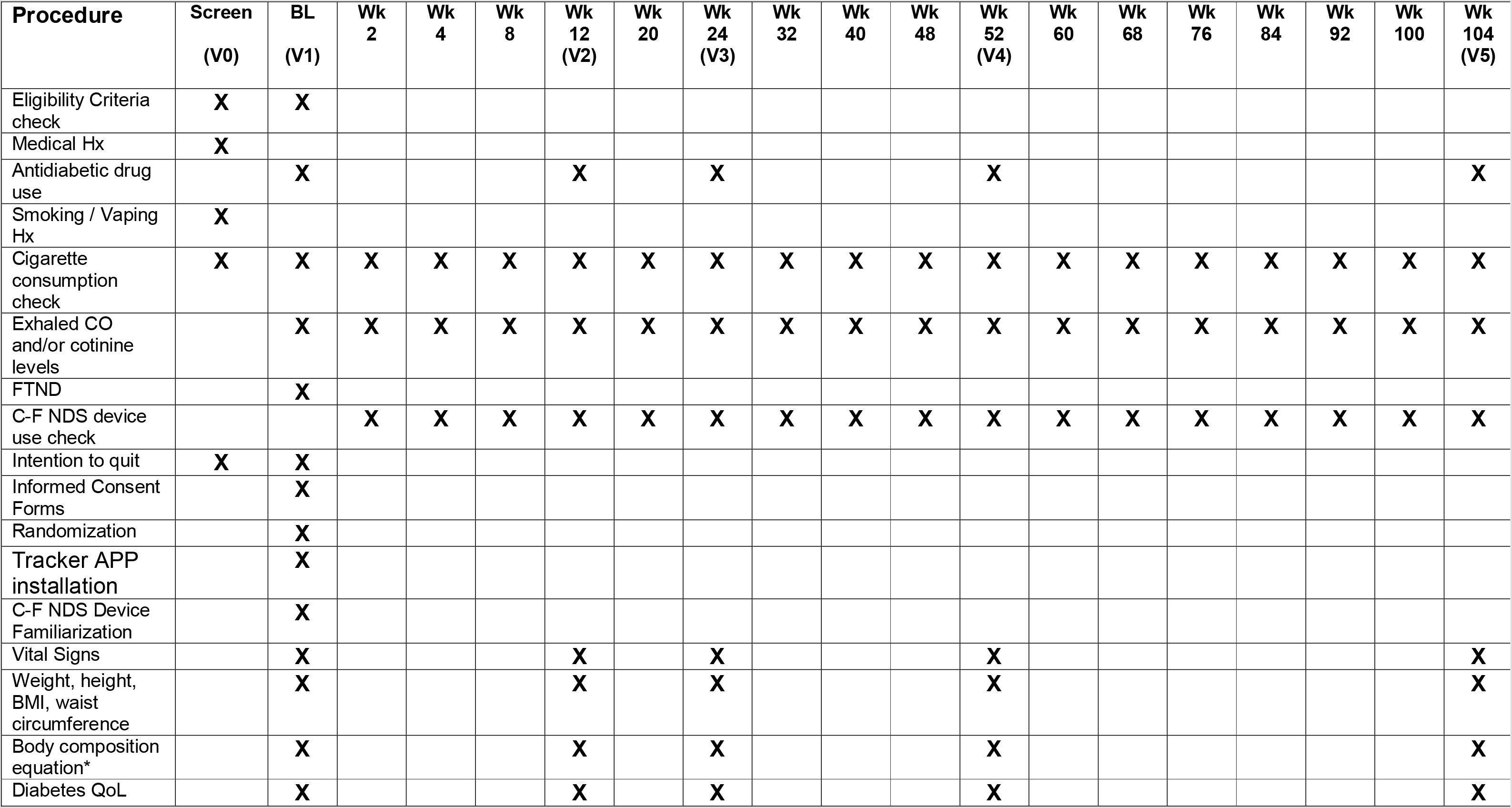
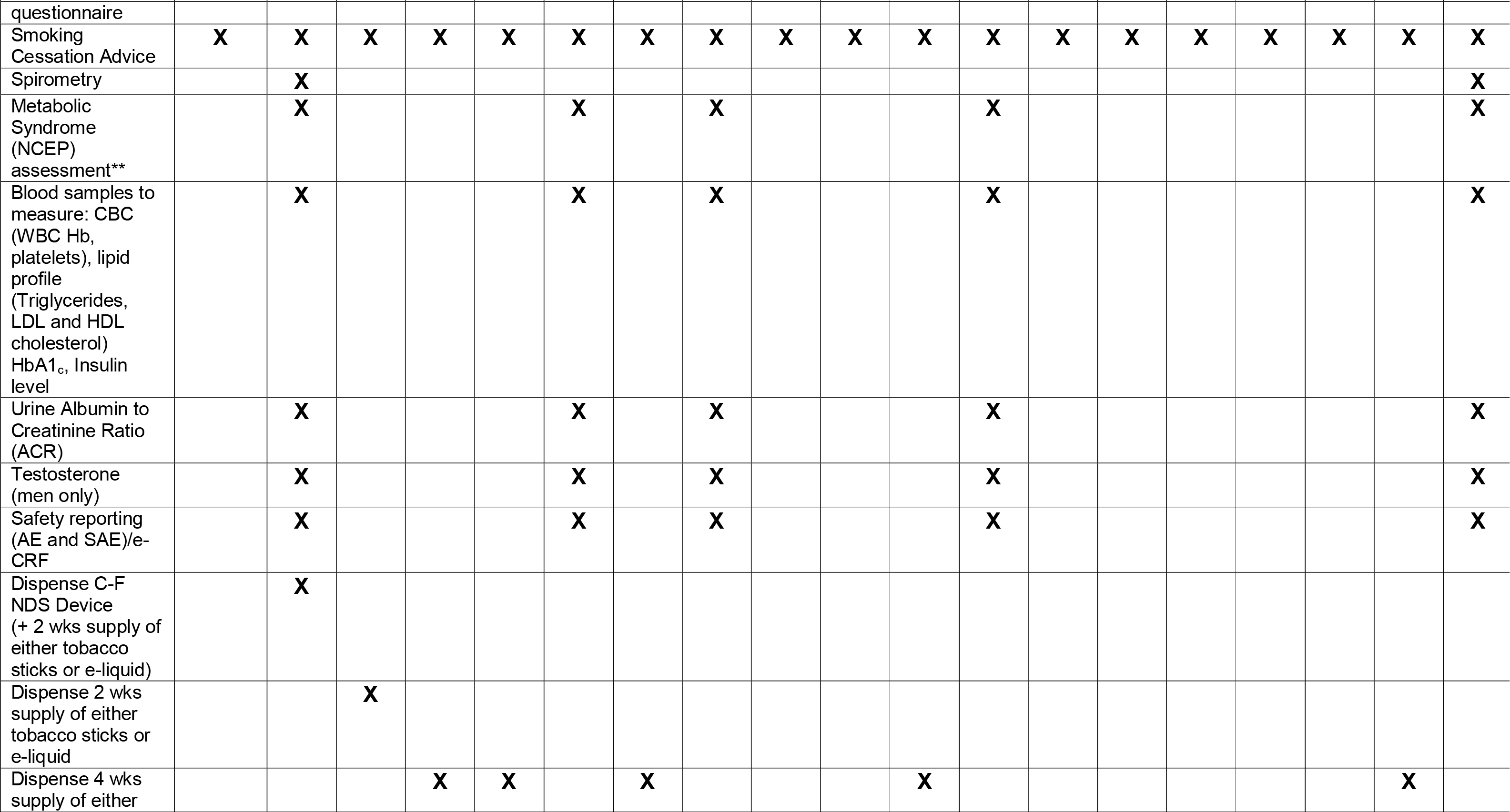
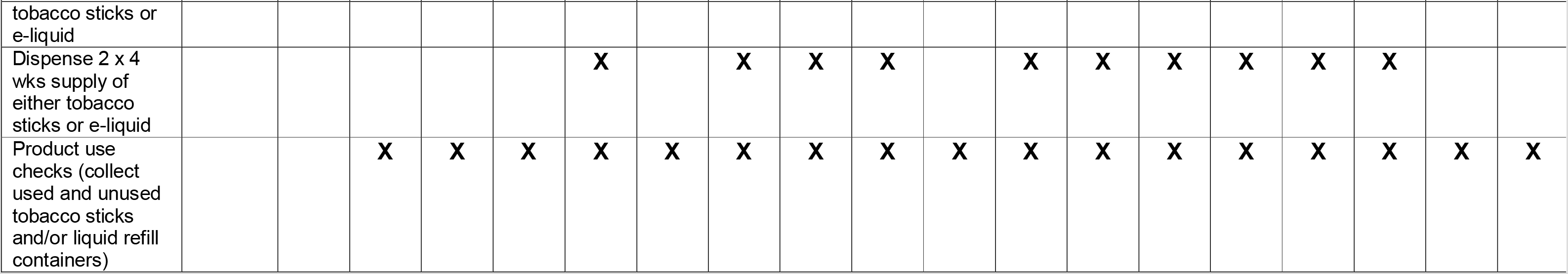
STUDY ASSESSMENTS (Arm B – switch to C-F NDS)

For patients randomized into arm B, between those clinical visits, additional nonclinical visits aiming to replace the used consumables are planned. At non-clinical visits, study investigators will also have the opportunity to stimulate retention and check compliance. In order to perform an evaluation of the habitual pattern of use of the CF-NDS and to verify product adherence, patients randomized into arm B will return all empty, part-used, and unused consumables at each visit.

At each visit, all participants will be advised and encouraged to completely quit smoking (cigarette or C-F NDS). They will explicitly be told about the risks associated with smoking and at every contact time-point offered referral to local free cessation programs. Premature withdrawal from the study may occur if participants: 1) experiences a severe adverse event (SAE); 2) sustain any protocol deviations occur during the conduct of the study, which cannot be corrected; 3) is uncooperative, including non-attendance; 4) decide to stop his/her participation at any moment of the study; 5) becomes pregnant. The trial will formally end on the date of the last visit of the last patient in the last country undertaking the trial.

### Objectives and Endpoints

The primary objective of DIASMOKE is to assess the impact of sustained use of CF-NDS on the proportion of patients with Metabolic Syndrome, as defined by National Cholesterol Education Program (NCEP) MetS score (21) below the diagnostic threshold (< 3).The primary outcome of the study will be change in prevalence of an NCEP MetS score < 3 between baseline and 2 years follow-up, with comparison being made between T2DM patients randomized to each arm of the study.

Change in prevalence will also be assessed at 3 months, 6 months, and 1 year, as secondary outcomes. All assessments at each time-point will be undertaken in all participants in both arms. Considering the results of a number of lifestyle modification interventions, the absolute reduction in MetS prevalence following substantial smoking cessation is expected to be no less than 15% [22–26].

The main prespecified secondary endpoint is an absolute change in the sum of the individual factors of the Metabolic Syndrome (as defined by NCEP criteria) measured at each study time-point (between and within study groups). Other secondary endpoints include change in each individual factor of the Metabolic Syndrome (as defined by NCEP criteria) measured at each study time-point (between and within study groups) and change of the following variables measured at each study time-point (between and within study groups).

### Statistical Considerations

#### Powering and Sample Size Calculation

For this study, the following input assumptions were considered:

- The absolute reduction in MetS prevalence following substantial smoking cessation is expected to be 15%, based on the results of a range of lifestyle modification interventions (22–26)
- The baseline prevalence of MetS in T2DM is expected to be 70% (27–30)

Sample size was calculated on the basis of demonstration of superiority, using an assumption of normal distribution, as described by Pocock (31). Significance level was set at 5% (α = 0.05), with a power of 80% (β = 0.20). On this basis, the minimum number of patients with analysable data required is 160 per treatment arm (N).

Further assumptions at the planning stage included an estimated 50% proportion of patients randomized to CF-NDS who are expected to achieve sustained reduction in cigarette consumption of at least 80% for the duration of the study (%_SusRed_) (32–36).

The adjusted number of patients in the intervention arm (N_2_) was therefore increased to 320:

N_2_ = N/%_susRed_ = 320 (N_2_ indicating the final number of patients required after taking into consideration the 50% sustained reduction figure).

Additionally, the expected number of patients in both arms withdrawing from the trial over 2 years is estimated at 20% (37–39). The total number of patients recruited to each treatment arm was therefore increased by this amount:

Intervention arm: 320 × 1.2 = 384

Control arm: 160 × 1.2 = 192

Total patients both arms = 576

#### Statistical Analyses

The primary endpoint for the statistical analysis is defined as the between-groups difference in calculated prevalence of MetS after at least 24 months of follow-up. The Full Analysis Set (FAS) comprises all patients randomized to the intervention arm who achieve a sustained reduction in cigarette consumption of at least 80% across the full duration of follow-up combined with all patients randomized into the standard care control group. The FAS will be the primary analysis set for all efficacy analysis. Two approaches to the primary analysis will be used:

a. Unadjusted analysis, based on a direct comparison of the change in prevalence. Z test will be used to assess the significance of difference between the two groups in the prevalence percentage changes from baseline to 24-month visit.
b. Adjusted analysis. Baseline demographics, clinical and concomitant therapeutic characteristics will be analysed to identify potential confounders for the primary outcome that are unbalanced between treatment groups. The primary outcome will then be re-analysed using a generalised linear model adjusting for all identified confounders.

Any difference between groups will be assessed for statistical significance at a 2-sided alpha of 0.05.

## RESULTS

Patient recruitment will start in October 2020 and enrolment is expected to be completed by August 2021. Results will be reported between 2023 and in 2024.

## DISCUSSION

Little is known about the impact of combustion-free nicotine delivery systems (C-F NDS) on T2DM patients who smoke. Products that do not require combustion to deliver nicotine, such as e-cigarettes (ECs) and heated tobacco products (HTPs) are substituting conventional cigarettes globally (17). They potentially offer substantial reduction in exposure to harmful and potentially harmful chemical constituents compared to conventional cigarettes (18–20, 40–42). DIASMOKE will be the first study determining the overall health impact of using such technologies in diabetic patients. Undoubtedly, it is desirable for patients to avoid consumption of any tobacco related inhalation products, but in order for Governments and clinicians to provide guidance about cigarette substitution, robust evidence-based information is required.

We designed this international RCT to gather such evidence. In particular, we will be testing the hypothesis that avoiding exposure to cigarette smoke toxicants may translate to measurable improvement in cardiovascular risk factors when T2DM patients who smoke switch to using C-F NDS compared with T2DM patients who continue to smoke conventional cigarettes. Several parameters measured in this study are associated with the development cardiovascular diseases (such as high blood pressure, elevated blood cholesterol, and BMI > 25) and some of these indicators have been shown to improve relatively soon after smoking cessation [43–45]. Consequently, the profile of these changes after switching to CF-NDS could provide valuable insights into the overall potential of CF-NDS to reduce the risk of cardiovascular disease.

The decision for a switching study design in DIASMOKE has been guided by the notion that C-F NDS have been promoted as substitutes for tobacco cigarettes. In a switching study of smokers the reference product is their own brand tobacco cigarette. The length of the study was based on the consideration that changes in the primary endpoint could be reasonably observed as early as 6-months. It is however possible that a much longer follow-up period could be necessary to firmly establish findings consistency over time, hence study duration was extended to 24-months. The RCT study design will provide a robust answer to determine the health impact of C-F NDS use on diabetic patients. Clearly, randomization will equalize variation in smoking history and other variables between study arms, thus ensuring high quality data. Importantly, the entire study is designed keeping the welfare of all participants at its centre; at every contact smokers will be asked to stop all types of smoking and provided with free local referrals for cessation smoking programs.

Compliance to the study protocol is critical as failure to fully or largely replace conventional cigarettes with C-F NDS would reduce or nullify the expected changes in study endpoints. Participants will be reminded on the importance of adhering to their randomised product allocation and of abstaining from or greatly reducing conventional cigarette consumption (by at least 80% from their baseline value of cigarette smoked in a day) at every contact. They will also be informed that biochemical verification of compliance as well as assessments of adherence will be conducted at each clinic visit. In addition, any noncompliance will be recorded in the study diary after counting all empty, part-used and unused consumables returned at each visit, and tracked by the APP. Although not expected that compliance for this study will be materially different compared to other comparable studies, our power calculations are over-estimated to take account of a non-compliance rate of 50%. Thus, the C-F NDS population will be oversampled by adopting a 1:2 randomization ratio scheme (i.e. for every patient randomized in the control population, two will be randomized in the C-F NDS population). Lastly, trial attendance and retention of the C-F NDS population will also be improved by asking participants to return to the clinic for their regular re-supply of tobacco sticks/e-cigarette cartridges/e-liquids refill bottles.

This study has several innovative features. To improve adherence to C-F NDS (and maximize overall compliance to the study protocol), patients randomized to switching to C-F NDS use will be offered a wide selection of different products (reflecting the most popular of those commercially available in each participating country) in order to choose the C-F NDS of their preference. Given that the population sample in DIASMOKE is mostly made of elderly patients, we will only offer devices that can ensure a likely user-friendly experience (i.e. easy to refill PODs, prefilled PODs, and heated tobacco devices). We expect that when participants are freely provided C-F NDS of their choosing they will be more likely to adopt the new technology and switch away from their own conventional cigarettes. Moreover, the study findings will not be product specific and unlikely to be limited in generalizability.

Substantiation of the reduced risk potential of long-term C-F NDS use is virtually unexplored. Data from DIASMOKE will be an important addition to the growing body of evidence in the field of understanding the health impact of combustion-free nicotine delivery technologies and will provide valuable insights into the overall potential of these products to reduce the risk of cardiovascular disease in individuals, particularly diabetic patients.

## Data Availability

The data that support the findings of this study are available from the corresponding author, PS, upon reasonable request.

